# A feasibility study of perioperative vitamin D supplementation in patients undergoing colorectal cancer resection

**DOI:** 10.1101/2022.08.18.22278022

**Authors:** P.G. Vaughan-Shaw, L.F. Buijs, J.P. Blackmur, A. Ewing, E. Theodoratou, Li-Yin. Ooi, F.V.N. Din, S.M. Farrington, M.G. Dunlop

**Affiliations:** MRC Human Genetics Unit, Institute of Genetics and Cancer, University of Edinburgh, Edinburgh, United Kingdom; Cancer Research UK Edinburgh Centre, Institute of Genetics and Cancer, University of Edinburgh, Edinburgh, United Kingdom; Centre for Global Health Research, Usher Institute for Population Health Sciences and Informatics, University of Edinburgh, Edinburgh, United Kingdom; Department of Pathology, National University Hospital, Singapore

## Abstract

**BACKGROUND:** Post-operative vitamin D supplementation improves CRC survival outcomes in randomised trials. The aim of this study was to test the feasibility, safety and efficacy of vitamin D in the pre- and perioperative period in patients undergoing CRC surgery.

**METHODS:** Patients were given 3200IU oral cholecalciferol per day perioperatively. Serial serum 25OHD was measured by liquid chromatography tandem mass spectrometry and compared to untreated CRC controls. 25OHD/CRP levels were compared using adjusted generalized linear mixed-effects models.

**RESULTS:** 122 patients underwent serial perioperative sampling, including 41 patients given high-dose perioperative supplementation. 25OHD was higher in patients given high-dose supplementation compared to controls at all peri-operative timepoints (P<0.001) and supplementation attenuated the post-operative drop in 25OHD (46% vs 24% drop). Rate of vitamin D insufficiency was significantly less in those on supplementation (14% *vs*. 56%, P=0.003), with multivariate modelling indicating a ∼60nmol/l higher 25OHD compared to control patients (P=1.22E-15). Supplementation was associated with lower peri-operative CRP (P=0.035).

**DISCUSSIONS:** High dose pre-operative vitamin D supplementation is associated with higher perioperative 25OHD levels, lower rates of vitamin D insufficiency and reduced early postoperative CRP. These findings provide compelling rationale for early initiation of vitamin D supplementation after a diagnosis of CRC.

## INTRODUCTION

Despite substantial improvement in mortality from colorectal cancer (CRC) over recent years, ∼16,600 people die from CRC each year in the UK ^1^. Whilst surgery remains the mainstay of therapy, new therapeutic approaches are required. Compelling evidence from observational studies ^2^ demonstrate a link between low vitamin D and poor outcomes following a diagnosis of colorectal cancer ^3-8^, while a recent meta-analysis of trial data demonstrating a 30% reduction in adverse CRC outcomes with supplementation ^9^. Plasma 25-hydroxyvitamin D (25OHD) levels in CRC patients are consistently lower than in population controls ^10^, yet a key unanswered question is whether supplementation could provide benefit from the time of diagnosis. Abdominal surgery is a major physiological insult and 25OHD (the best marker and storage form of vitamin D) falls dramatically following bowel cancer surgery ^2^. 25OHD is also known to decrease following orthopaedic, cardiac and gynaecological surgery ^11-16^. Several explanations are proposed to explain the observed decrease, including circulatory fluid changes i.e. haemodilution ^14,17^ and/or systemic inflammatory responses to surgery. It follows that because 25OHD depletion is associated with adverse survival outcomes and supplementation can improve survival, studies to define when to initiate supplementation are required. While no study to date has assessed the impact of vitamin D supplements in patients undergoing colorectal cancer resection, cholecalciferol (vitamin D3) supplements have been shown to significantly improve 25OHD levels in patients with a historical diagnosis of CRC ^18^ and in patients undergoing chemotherapy for CRC ^19^.

Here, we have investigated the perioperative temporal variation in plasma 25OHD and CRP by serially sampling patients undergoing CRC resectional surgery and aimed to demonstrate that vitamin D supplementation is feasible, safe and effective in the perioperative period. These data will be essential in informing the design of future randomised trials of vitamin D and survival outcomes after colorectal cancer surgery.

## METHODS

The Study of Colorectal Cancer in Scotland (SOCCS) study is a national population-based case-control study designed to identify genetic and environmental factors that have an impact on CRC risk and survival outcome ^20^. The Scottish Vitamin D study (SCOVIDS) is a single-centre study designed to investigate the link between vitamin D, CRC risk and survival. Participants provided informed written consent, and research was approved by local research ethics committees (SOCCS 11/SS/0109 and 01/0/05; SCOVIDS 13/SS/0248) and National Health Service management (SOCCS 2013/0014, 2003/W/GEN/05; SCOVIDS 2014/0058) and registered with clinicaltrials.gov (NCT05506696). Clinical and sampling variables were collected from patient case notes and pathology records, entered into a prospective study database and extracted for analysis. We used data from patients recruited between 2012 and 2013 who underwent serial sampling during the preoperative and immediate postoperative period, as previously described ^2^. We then recruited patients to undergo high dose pre- and perioperative oral vitamin D supplementation (3200IU/ day cholecalciferol, Fultium-D3, Internis Pharmaceuticals Ltd, Huddersfield, UK), alongside a contemporaneous control group. All patients eligible for supplementation but not already established on vitamin D or multivitamin supplementation were considered for inclusion (Supplementary Tabble 1). Whether the patient was recruited by our research nurse, or surgical research fellows (PVS/ LB) determined whether they were allocated to the control or supplementation group respectively, itself determined by timetabling and clinic availability of each recruiter. Patients administered supplementation took 3200IU everyday preoperatively, including on the morning of surgery, and in the early postoperative period except where postoperative ileus occurred. Given the pragmatic methodology, no alteration to the normal patient pathway occurred and there was marked variation in duration of preoperative supplementation between patients, in part due to delays and disruption from the COVID-19 pandemic. Recruitment was paused during the first wave of the COVID-19 pandemic, significantly impacting on total study recruitment, while restrictions on hospital appointments and non-clinical sampling limited the number of post-discharge samples that could be taken.

### Plasma vitamin D assay

Samples were taken at preoperative assessment clinic and/or the day of surgery, and postoperatively (on the ward at 1-2 days, 3-5 days, 6-9 days) and at the first postoperative clinic appointment. COVID-19 restrictions and the emergency of telephone follow-up significantly impacted the ability to perform post-discharge sampling.

Plasma was extracted from blood sampled by venepuncture from a peripheral arm vein. To minimise any potential analytical variation in plasma 25OHD measurement, all samples were assayed in a single United Kingdom Accreditation Service (UKAS) accredited laboratory using a method traceable to National Standards of Science and Technology standard reference material. 21 Total 25OHD (25OHD2 and 25OHD3) was measured by liquid chromatography tandem mass spectrometry and analysed using the Waters® Xevo® TQ-S system (Waters Limited, Wilmslow, UK).

### Plasma CRP assay

To assess for potential relevance of the systemic inflammatory response, samples were assayed for CRP in a NHS Biochemistry Laboratory serving our hospital. CRP was measured using the Abbott Architect C series clinical chemistry analyser to standard sensitivity protocol, with the range of output values 0.2-480 mg/l. CRP internal quality control was performed daily, with 3 control samples assayed twice per day. Target mean of the three control samples was 3.2, 8.3 and 27.5mg/l with actual observed mean over 6 months of 3.2, 8.4 and 27.8mg/l respectively from 5151 completed assays. Coefficients of variation were 3.42%, 2.34% and 2.05% respectively.

### Sample size considerations

Data from our previously published control group ^2^ was used to inform a power calculation, with the current study design powered to identify a 50% suppression of the day 1-2 postoperative drop in 25OHD with supplementation (α=0.05 [one-tailed]; β=0.10, calculated sample size in each arm = 54).

### Patient and public involvement

This project is relevant to biomarker prediction and risk stratification and was assigned high priority in the seminal Association of Coloproctology of Great Britain and Ireland (ACPGBI) Delphi process ^22^. After draft protocol design, we developed our lay summary and PPI questionnaire in collaboration with the University of Edinburgh Patient Advisory Group and Patient Liaison Group of the ACPGBI. Thereafter we surveyed eight patients with colorectal cancer to assess the acceptability of the proposed serial sampling study and help inform our final study design.

### Patient, Tumour and Treatment-Related Variables

We considered and adjusted for patient-related factors previously established to influence 25OHD levels (age, sex, body mass index (BMI). ^23-25^ Tumour and treatment-related variables were collected to investigate putative effect on 25OHD, including American Joint Committee on Cancer (AJCC) stage. Information on tumour site, multiplicity and clinico-pathologic staging were obtained from clinical records, along with preoperative imaging. By using collated pathology, imaging, and clinical data; tumour stage was mapped onto the AJCC staging system (AJCC stage I to IV).

### Statistical analysis

All statistical analysis was undertaken in R (version 4.1.0) [26]. To account for seasonal differences in vitamin D status, reported 25OHD level was May-standardized by adjusting for sampling month using differences in age- and sex-adjusted monthly averages generated from SOCCS control data [27]. A linear mixed-effects model was applied to the serially sampled data to examine the association between perioperative changes in CRP and 25OHD (log-transformed) and treatment group. This multivariable model adjusted for age, gender, BMI and AJCC tumour stage. The package ‘*emmeans’* was used to compute contrasts between estimated marginal means to evaluate potential differences in 25OHD between treatment groups at each timepoint. Finally, a binomial generalized linear mixed-effects model was applied to test for differences in rate of vitamin D insufficiency between treatment groups and at each peri-operative timepoint, adjusting for age, gender, BMI and AJCC tumour stage.

## RESULTS

### Baseline demographics

We recruited a total of 122 patients who underwent serial perioperative sampling at up to 6 time-points, including 41 patients who received high-dose perioperative vitamin D supplementation (Table 1). No significant differences in baseline clinicopathological variables between the groups were observed. No differences between the previously published control group and newly recruited control group were observed, including in baseline 25OHD (46.3 *vs*. 47.6nmol/l, P=0.86; Supplementary Tabble 2).

**TABLE 1.**
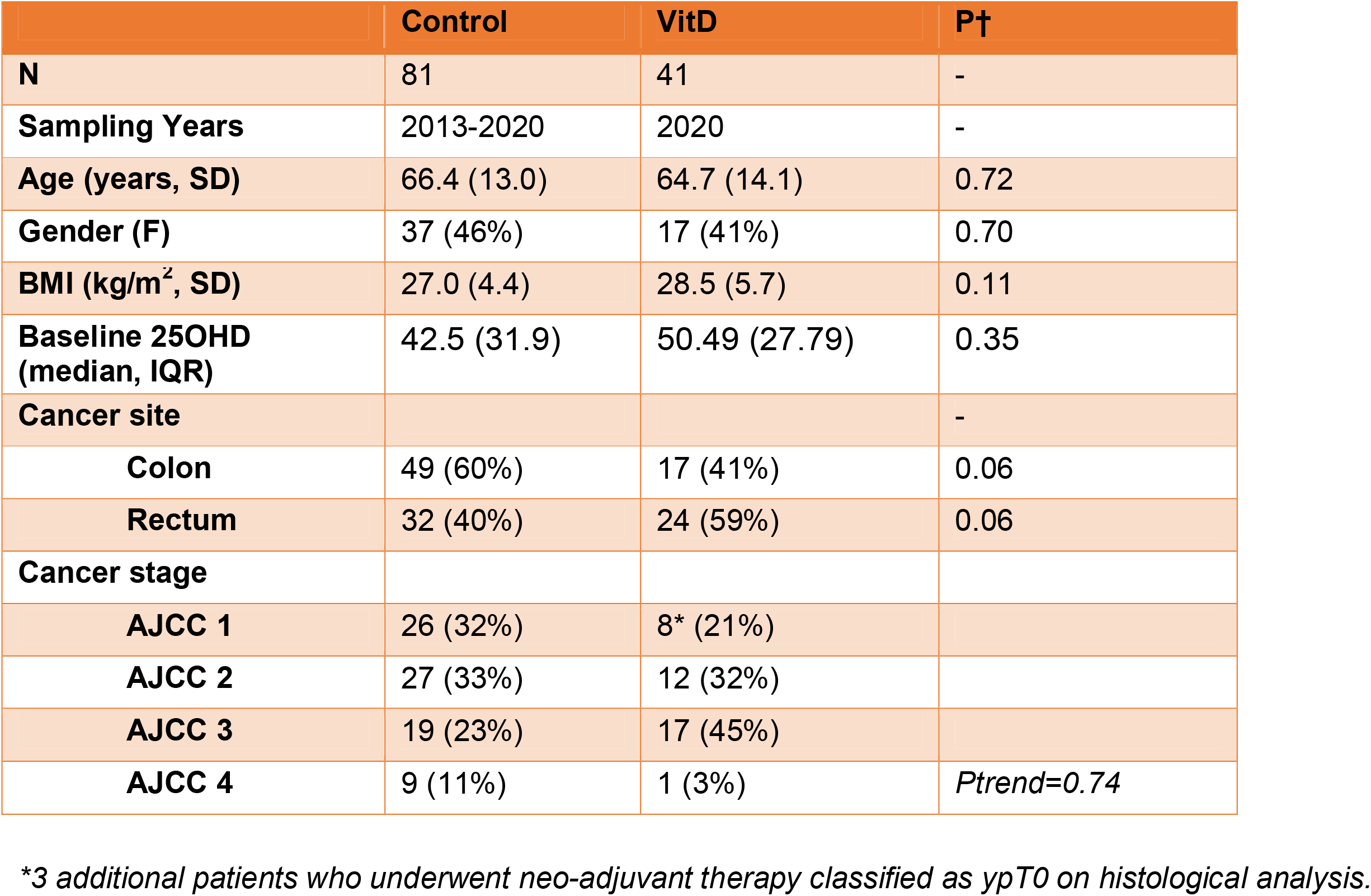
Clinicopathological demographics of included patients.

### Efficacy and safety of perioperative supplementation

No serious adverse events occurred in those taking perioperative supplementation, which was well tolerated in study participants. In patients with both a pre- and post-supplementation sample (N=13), supplementation induced a 2.1 fold-increase (48.2nmol/; 95%CI 20.7-75.6; P=0.01) in 25OHD level (Table 2, Figure 1). Overall, patients took preoperative supplementation for median 25 days (range 9-84) with no correlation between number of days’ supplementation and either increase in 25OHD (R=-0.20, P=0.5) or 25OHD level on day of surgery (R=0.14, P=0.39). Dose diaries assessed for 8 subjects, indicated high compliance with daily supplementation (316 out of 319 days perioperative supplementation completed).

**TABLE 2.**
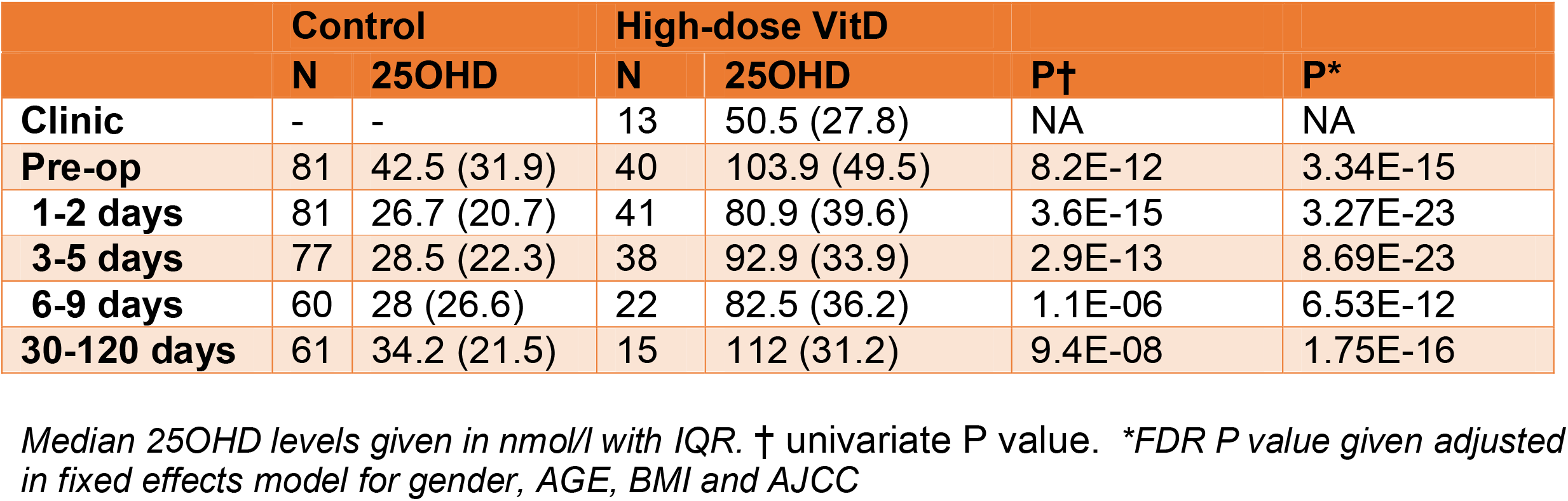
May-adjusted perioperative 25OHD.

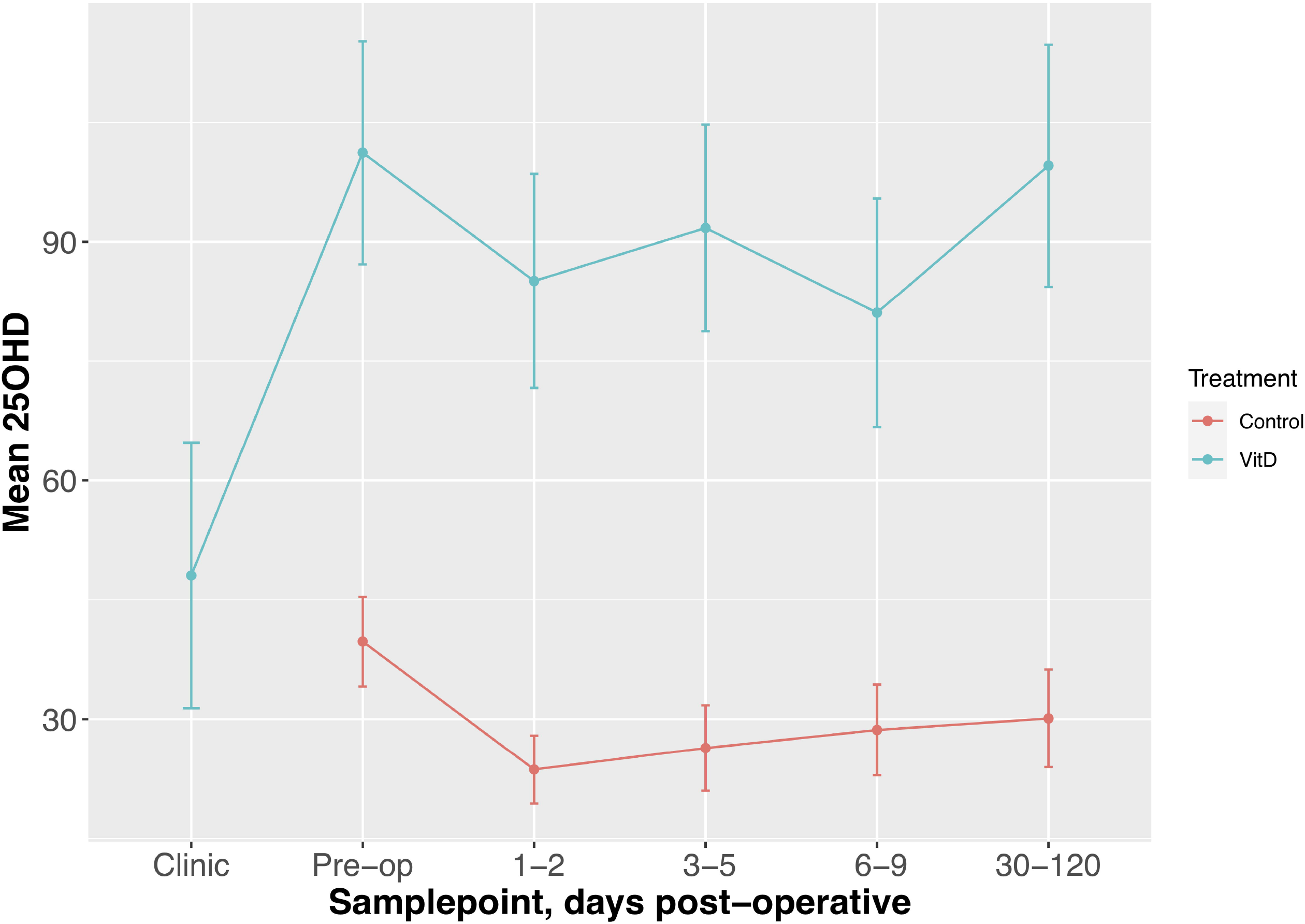

Supplemented patients had higher 25OHD levels on the day of surgery (P=8.2E-12, Table 2) and in the early postoperative period when compared to control patients (e.g. 1-2 days 80.9nmol/l *vs*. 26.7nmol/l, P=3.6E-15; Table 2, Figure 1). Crucially, the multivariate fixed-effects model confirmed a significant and clinically relevant impact of supplementation on perioperative 25OHD levels, after adjusting for age, gender, BMI and AJCC stage (Table 2; Supplementary Tabble 3). Supplementation also reduced the prevalence of perioperative vitamin D insufficiency (25OHD<50nmol/l), with 14% of supplemented patients having early postoperative insufficiency compared to 56% of control patients (P=0.003, Table 4). Finally, although 25OHD levels dropped in all treatment groups following surgery, supplementation attenuated the early post-operative drop (e.g. 46% drop *vs*. 24% drop at days 1-2, P=0.0003). (Figure 1; Supplementary Tabble 4).

### Impact of supplementation on postoperative CRP levels

CRP levels assayed at the same time as 25OHD were compared across treatment groups. As expected, CRP increased postoperatively in all groups, yet the increase appeared less in those taking high-dose supplementation, with lower early postoperative CRP levels seen in these patients (e.g. 3-5 days 80.5mg/l *vs*. 129mg/l, P=0.04, Table 5, Supplementary Figure 2). The multivariate mixed-effects model indicated that supplementation had a modest impact on peri-operative CRP levels after adjusting for age, gender, BMI and AJCC stage (P=0.035; Table 3; Supplementary Tabble 5).

**TABLE 3.**
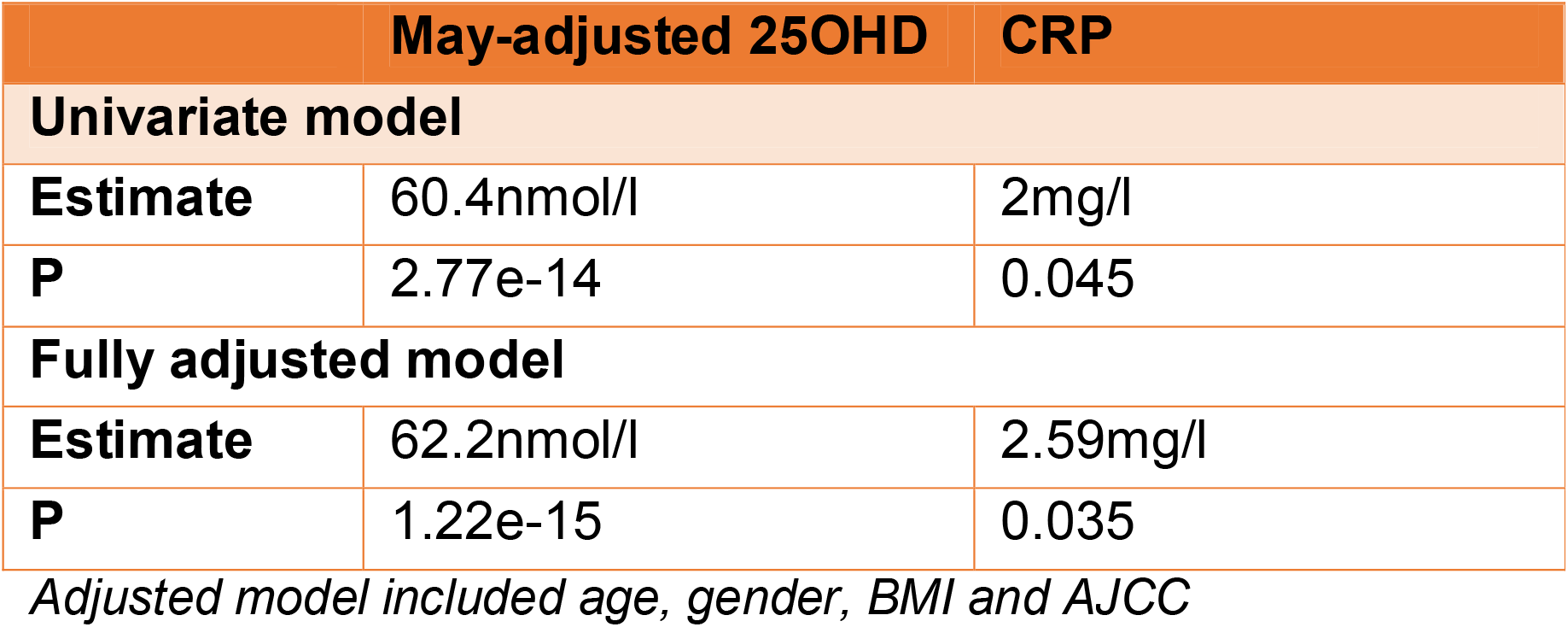
Impact of treatment on 25OHD and CRP levels in mixed-effects model.

**TABLE 4.**
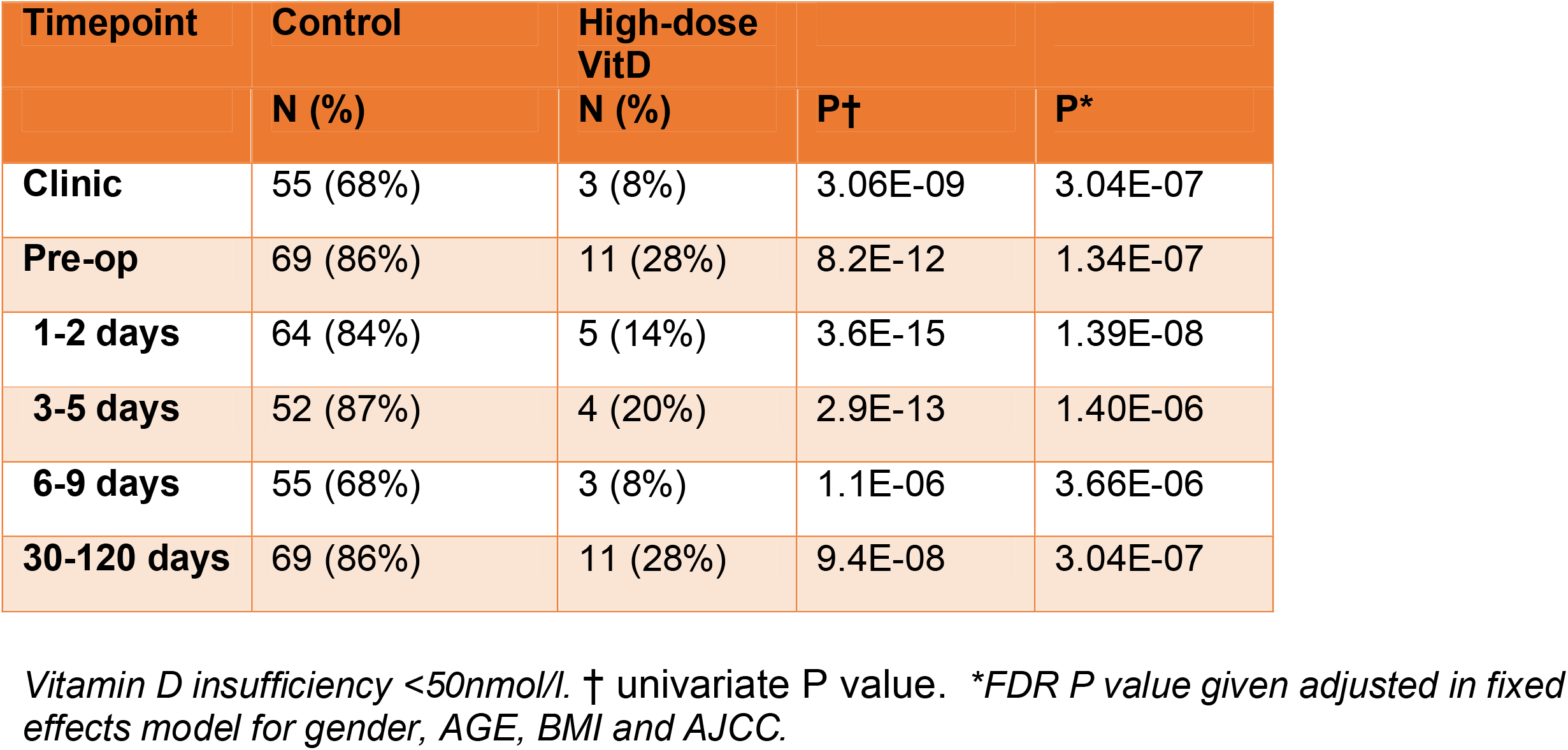
Perioperative Vitamin D.

**TABLE 5.**
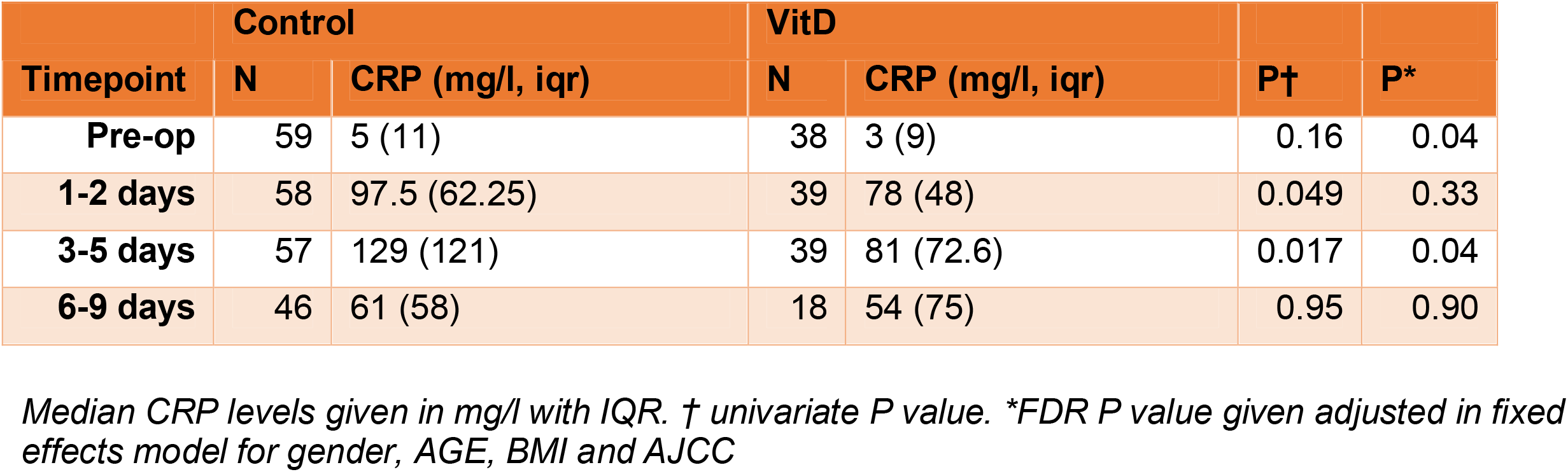
Perioperative CRP levels in control and supplemented patients.

## DISCUSSION

High-dose perioperative vitamin D supplementation is safe and well tolerated in patients undergoing colorectal cancer surgery. Compared to control patients, supplementation induces a significant increase in perioperative 25OHD levels, a smaller relative drop in 25OHD following surgery and lower rates of postoperative vitamin D insufficiency. Meanwhile, early postoperative CRP appears to lower in patients on supplementation supporting the role of vitamin D in regulation of inflammatory processes.

This study provides evidence for a beneficial effect of vitamin D supplementation on perioperative vitamin D status while a recent meta-analysis of RCT data reported a significant reduction in CRC mortality with vitamin D supplementation ^9,26^. Taken together, these data support early initiation of supplementation at the point of CRC diagnosis. While vitamin D repletion has already been shown to be feasible in patients undergoing chemotherapy ^3,4,19^, no study to date has explored its use in CRC patients in the perioperative period. Vitamin D levels are known to drop following surgery ^2^, which is confirmed here in all treatment groups. Here, we demonstrate that despite the insult of resectional cancer surgery, and associated changes in gut motility and absorption, perioperative supplementation induces a marked increase in preoperative 25OHD levels and attenuates the drop in 25OHD following surgery. Given the tight physiological autoregulation that exists, it is unlikely that mechanisms that confer the observed beneficial impact of vitamin D on CRC survival occur in a linear manner, but rather that a threshold effect exists. Given that our previous work suggests that a 25OHD threshold of ∼45-50nmol/l appears to most strongly associate with survival, it is relevant that in the current study supplementation significantly reduced postoperative vitamin D insufficiency (25OHD<50nmol/l).

To date, no trial has investigated the optimum time to initiate supplementation. Indeed, few of the supplementation trials in CRC patients include patients undergoing curative resection, with one such trial (AMATERASU trial), recruiting patients at the first outpatient visit after surgery ^5^. It is relevant that in population trials where recruitment and supplementation occur before the diagnosis of incident cases of CRC, a beneficial effect of vitamin D supplementation on survival is still seen. Furthermore, 25OHD levels sampled preoperatively and at the earliest postoperative timepoint (<6 months) are already associated with survival in observational data ^2^. Therefore, given that this cheap supplement has now been shown to be safe and well tolerated in the perioperative period, there is compelling rationale to start at the point of diagnosis, especially given the increasing use of neo-adjuvant therapy which can delay resectional CRC surgery for many months.

Previous pre-clinical and human intervention studies provide clues to the mechanisms that may underlie the beneficial effect of supplementation on CRC survival and provide no contra-indication to earlier supplementation. 1,25-dihydroxyvitamin D3 modulates immune and inflammatory pathway genes in large bowel epithelium ^27^ while oral supplementation induces transcriptomic changes in rectal mucosa that are linked to anti-tumour effects ^28^. Vitamin D also regulates multiple inflammatory processes both *in vitro* and *in vivo*, including those involved in CRC such as oxidative stress and the cyclooxygenase and NF-kB pathways ^29,30^. Effects on inflammatory processes are consistent with the lower postoperative CRP levels in supplemented patients in the current study, supporting the notion that vitamin D might be causally implicated in the SIR response following surgery. This provides further potential mechanism to improved survival outcomes with supplementation given that CRP, an established marker of inflammation, is strongly correlated with CRC survival ^31,32^.

The current study has a number of limitations. We acknowledge that supplemented patients recruited in 2020 were compared against a combined control group including patients recruited in 2013. As such, unmeasured historical differences in demographics, genetic background (e.g. vitamin D receptor or pathway SNPs ^33^) or clinical factors may confound the observed effect of supplementation. However, we observed no difference in baseline or peri-operative 25OHD between the two control groups. Furthermore, mixed-modelling identified a significant association between supplementation and increased 25OHD level when comparing supplemented patients with the newest control cohort (∼60nmol/l; P=3.51E-08). Nevertheless, we cannot fully exclude the possibility that differences in perioperative 25OHD levels are due to factors other than supplementation (i.e. confounding factors) because this was not a randomised study. Third, the power of our study was limited by COVID-19 which markedly impacted total study recruitment and sampling. Finally, while our data supports the early initiation of supplementation in patients undergoing CRC surgery given its effects on 25OHD level and extrapolating from previous trial evidence of survival benefit, we do not provide direct evidence of survival benefit from early supplementation.

In conclusion, we report for the first time on the feasibility and safety of perioperative vitamin D supplementation in patients undergoing colorectal cancer surgery. We identified a positive effect of supplementation on perioperative 25OHD levels with lower rates of postoperative vitamin D insufficiency and reduced early postoperative CRP. Our findings provide compelling rationale for early initiation of vitamin D supplementation after a diagnosis of CRC. Future randomised trials of supplementation with a defined endpoint of a beneficial effect on survival outcomes should consider supplementation from the point of diagnosis.

## Data Availability

All data produced in the present study are available upon reasonable request to the authors

## Abbreviations

25OHD: 25-hydroxyvitamin D
AJCC: American Joint Committee on Cancer stage
BMI: Body mass index
CI: Confidence interval
CRC: Colorectal cancer
CRP: C-reactive protein
SIR: Systemic inflammatory response
SCOVIDS: Scottish Vitamin D study
SOCCS: Study of Colorectal Cancer in Scotland

## ADDITIONAL INFORMATION

## Acknowledgements

We acknowledge the excellent technical support from Marion Walker and Stuart Reid. We are grateful to Donna Markie, and all those who continue to contribute to recruitment, data collection, and data curation for the Study of Colorectal Cancer in Scotland studies. We acknowledge the nursing and study facilities provided by the Edinburgh Wellcome Trust Clinical Research Facility. Finally, we acknowledge that these studies would not be possible without the patients and surgeons who take part. The manuscript is available on medRxiv, MEDRXIV/2022/278022

## Authors’ contributions

Study concepts: PVS, LFB, JPB, LYO, FVND, SMF, MGD

Study design: PVS, LFB, LYO, JPB, ET, SMF, MGD

Data acquisition: PVS, LYO, LFB, JPB,

Quality control of data and algorithms: PVS, AE

Data analysis and interpretation: PVS, AE

Statistical analysis: PVS, AE

Manuscript preparation: PVS, LFB, JPB, AE, MT, SMF, MGD

Manuscript editing: PVS, LFB, JPB, AE, MT, SMF, MGD

Manuscript review: PVS, ET, AE, FVND, SMF, MGD

Funding acquisition: PVS, SMF, MGD

Project administration: MGD

Supervision: MGD, SMF

## Ethics approval and consent to participate

Participants provided informed written consent, and research was approved by local research ethics committees (SOCCS 11/SS/0109 and 01/0/05; SCOVIDS 13/SS/0248) and National Health Service management (SOCCS 2013/0014, 2003/W/GEN/05; SCOVIDS 2014/0058).

## Consent for publication

Not applicable

## Data Availability Statement

Data available upon reasonable request

## References

1. CRUK. Cancer statistics. https://www.cancerresearchuk.org/health-professional/cancer-statistics/statistics-by-cancer-type. Accessed 24/12/2019, 2019 (2019).

2. Vaughan-Shaw PG, Zgaga L, Ooi LY, Theodoratou E, Timofeeva M, Svinti V et al. Low plasma vitamin D is associated with adverse colorectal cancer survival after surgical resection, independent of systemic inflammatory response. Gut 2020; 69(1): 103-111; e-pub ahead of print 2019/04/27; doi 10.1136/gutjnl-2018-317922.

3. Antunac Golubic Z, Barsic I, Librenjak N, Plestina S. Vitamin D Supplementation and Survival in Metastatic Colorectal Cancer. Nutr Cancer 2018; 70(3): 413-417; e-pub ahead of print 2018/03/14; doi 10.1080/01635581.2018.1445766.

4. Ng K, Nimeiri HS, McCleary NJ, Abrams TA, Yurgelun MB, Cleary JM et al. Effect of High-Dose vs Standard-Dose Vitamin D3 Supplementation on Progression-Free Survival Among Patients With Advanced or Metastatic Colorectal Cancer: The SUNSHINE Randomized Clinical Trial. JAMA 2019; 321(14): 1370-1379; e-pub ahead of print 2019/04/10; doi 10.1001/jama.2019.2402.

5. Urashima M, Ohdaira H, Akutsu T, Okada S, Yoshida M, Kitajima M et al. Effect of Vitamin D Supplementation on Relapse-Free Survival Among Patients With Digestive Tract Cancers: The AMATERASU Randomized Clinical Trial. JAMA 2019; 321(14): 1361-1369; e-pub ahead of print 2019/04/10; doi 10.1001/jama.2019.2210.

6. Trivedi DP, Doll R, Khaw KT. Effect of four monthly oral vitamin D3 (cholecalciferol) supplementation on fractures and mortality in men and women living in the community: randomised double blind controlled trial. BMJ 2003; 326(7387): 469; e-pub ahead of print 2003/03/01; doi 10.1136/bmj.326.7387.469 326/7387/469 [pii].

7. Manson JE, Bassuk SS, Buring JE, Group VR. Principal results of the VITamin D and OmegA-3 TriaL (VITAL) and updated meta-analyses of relevant vitamin D trials. J Steroid Biochem Mol Biol 2019; 198: 105522; e-pub ahead of print 2019/11/17; doi 10.1016/j.jsbmb.2019.105522.

8. Wactawski-Wende J, Kotchen JM, Anderson GL, Assaf AR, Brunner RL, O’Sullivan MJ et al. Calcium plus vitamin D supplementation and the risk of colorectal cancer. N Engl J Med 2006; 354(7): 684-696; e-pub ahead of print 2006/02/17; doi 354/7/684[pii]10.1056/NEJMoa055222.

9. Vaughan-Shaw PG, Buijs LF, Blackmur JP, Theodoratou E, Zgaga L, Din FVN et al. The effect of vitamin D supplementation on survival in patients with colorectal cancer: systematic review and meta-analysis of randomised controlled trials. Br J Cancer 2020; e-pub ahead of print 2020/09/16; doi 10.1038/s41416-020-01060-8.

10. Theodoratou E, Palmer T, Zgaga L, Farrington SM, McKeigue P, Din FV et al. Instrumental variable estimation of the causal effect of plasma 25-hydroxy-vitamin D on colorectal cancer risk: a mendelian randomization analysis. PLoS One 2012; 7(6): e37662. e-pub ahead of print 2012/06/16; doi 10.1371/journal.pone.0037662 PONE-D-11-23990 [pii].

11. Skuladottir GV, Cohen A, Arnar DO, Hougaard DM, Torfason B, Palsson R et al. Plasma 25-hydroxyvitamin D2 and D3 levels and incidence of postoperative atrial fibrillation. J Nutr Sci 2016; 5: e10; doi 10.1017/jns.2015.38.

12. Pilka R, Marek R, Adam T, Kudela M, Ondrova D, Neubert D et al. Systemic Inflammatory Response After Open, Laparoscopic and Robotic Surgery in Endometrial Cancer Patients. Anticancer Res 2016; 36(6): 2909–2922.

13. Louw JA, Werbeck A, Louw ME, Kotze TJ, Cooper R, Labadarios D. Blood vitamin concentrations during the acute-phase response. Crit Care Med 1992; 20(7): 934–941.

14. McNally JD, Menon K, Chakraborty P, Fisher L, Williams KA, Al-Dirbashi OY et al. Impact of anesthesia and surgery for congenital heart disease on the vitamin d status of infants and children: a prospective longitudinal study. Anesthesiology 2013; 119(1): 71–80; doi 10.1097/ALN.0b013e31828ce817.

15. Reid D, Toole BJ, Knox S, Talwar D, Harten J, O’Reilly DS et al. The relation between acute changes in the systemic inflammatory response and plasma 25-hydroxyvitamin D concentrations after elective knee arthroplasty. Am J Clin Nutr 2011; 93(5): 1006–1011; doi 10.3945/ajcn.110.008490.

16. Waldron JL, Ashby HL, Cornes MP, Bechervaise J, Razavi C, Thomas OL et al. Vitamin D: a negative acute phase reactant. J Clin Pathol 2013; 66(7): 620–622; doi 10.1136/jclinpath-2012-201301.

17. Krishnan A, Ochola J, Mundy J, Jones M, Kruger P, Duncan E et al. Acute fluid shifts influence the assessment of serum vitamin D status in critically ill patients. Crit Care 2010; 14(6): R216; doi 10.1186/cc9341.

18. Vaughan-Shaw P.G. OLY, Timofeeva M., Svinti V., Walker M., Farrington S.M., Din F.V., Dunlop M.G. Peri-operative plasma vitamin D level in colorectal cancer patients and effect of vitamin D supplementation on colorectal cancer patients. Colorectal disease : the official journal of the Association of Coloproctology of Great Britain and Ireland 2016; 18(Supp 2): 34.

19. Savoie MB, Paciorek A, Zhang L, Van Blarigan EL, Sommovilla N, Abrams D et al. Vitamin D Levels in Patients with Colorectal Cancer Before and After Treatment Initiation. J Gastrointest Cancer 2019; 50(4): 769-779; e-pub ahead of print 2018/07/31; doi 10.1007/s12029-018-0147-7.

20. Theodoratou E, Kyle J, Cetnarskyj R, Farrington SM, Tenesa A, Barnetson R et al. Dietary flavonoids and the risk of colorectal cancer. Cancer Epidemiol Biomarkers Prev 2007; 16(4): 684-693; e-pub ahead of print 2007/04/10; doi 16/4/684[pii]10.1158/1055-9965.EPI-06-0785.

21. Knox S, Harris J, Calton L, Wallace AM. A simple automated solid-phase extraction procedure for measurement of 25-hydroxyvitamin D3 and D2 by liquid chromatography-tandem mass spectrometry. Ann Clin Biochem 2009; 46(Pt 3): 226–230; doi 10.1258/acb.2009.008206.

22. Tiernan J, Cook A, Geh I, George B, Magill L, Northover J et al. Use of a modified Delphi approach to develop research priorities for the association of coloproctology of Great Britain and Ireland. Colorectal Dis 2014; 16(12): 965-970; e-pub ahead of print 2014/10/07; doi 10.1111/codi.12790.

23. Kimlin MG, Lucas RM, Harrison SL, van der Mei I, Armstrong BK, Whiteman DC et al. The contributions of solar ultraviolet radiation exposure and other determinants to serum 25-hydroxyvitamin D concentrations in Australian adults: the AusD Study. Am J Epidemiol 2014; 179(7): 864-874; e-pub ahead of print 2014/02/28; doi kwt446[pii]10.1093/aje/kwt446.

24. Mithal A, Wahl DA, Bonjour JP, Burckhardt P, Dawson-Hughes B, Eisman JA et al. Global vitamin D status and determinants of hypovitaminosis D. Osteoporos Int 2009; 20(11): 1807–1820; doi 10.1007/s00198-009-0954-6.

25. Zgaga L, Theodoratou E, Farrington SM, Din FV, Ooi LY, Glodzik D et al. Plasma vitamin D concentration influences survival outcome after a diagnosis of colorectal cancer. J Clin Oncol 2014; 32(23): 2430–2439; doi 10.1200/JCO.2013.54.5947.

26. Keum N, Lee DH, Greenwood DC, Manson JE, Giovannucci E. Vitamin D supplementation and total cancer incidence and mortality: a meta-analysis of randomized controlled trials. Ann Oncol 2019; 30(5): 733-743; e-pub ahead of print 2019/02/24; doi 10.1093/annonc/mdz059.

27. Protiva P, Pendyala S, Nelson C, Augenlicht LH, Lipkin M, Holt PR. Calcium and 1,25-dihydroxyvitamin D3 modulate genes of immune and inflammatory pathways in the human colon: a human crossover trial. Am J Clin Nutr 2016; 103(5): 1224–1231; doi 10.3945/ajcn.114.105304.

28. Vaughan-Shaw PG, Grimes G, Blackmur JP, Timofeeva M, Walker M, Ooi LY et al. Oral vitamin D supplementation induces transcriptomic changes in rectal mucosa that are linked to anti-tumour effects. BMC Med 2021; 19(1): 174; e-pub ahead of print 2021/08/04; doi 10.1186/s12916-021-02044-y.

29. van Harten-Gerritsen AS, Balvers MG, Witkamp RF, Kampman E, van Duijnhoven FJ. Vitamin D, Inflammation, and Colorectal Cancer Progression: A Review of Mechanistic Studies and Future Directions for Epidemiological Studies. Cancer Epidemiol Biomarkers Prev 2015; 24(12): 1820–1828; doi 10.1158/1055-9965.EPI-15-0601.

30. Bostick RM. Effects of supplemental vitamin D and calcium on normal colon tissue and circulating biomarkers of risk for colorectal neoplasms. The Journal of steroid biochemistry and molecular biology 2015; 148: 86–95; doi 10.1016/j.jsbmb.2015.01.010.

31. McMillan DC, Canna K, McArdle CS. Systemic inflammatory response predicts survival following curative resection of colorectal cancer. Br J Surg 2003; 90(2): 215–219; doi 10.1002/bjs.4038.

32. McMillan DC. The systemic inflammation-based Glasgow Prognostic Score: a decade of experience in patients with cancer. Cancer Treat Rev 2013; 39(5): 534–540; doi 10.1016/j.ctrv.2012.08.003.

33. Revez JA, Lin T, Qiao Z, Xue A, Holtz Y, Zhu Z et al. Genome-wide association study identifies 143 loci associated with 25 hydroxyvitamin D concentration. Nat Commun 2020; 11(1): 1647; e-pub ahead of print 2020/04/04; doi 10.1038/s41467-020-15421-7.

