## Supplementar for "A feasibility study of perioperative vitamin D supplementation in patients undergoing colorectal cancer resection"

Supplementary Tables and Figures

**SUPPLEMENTARY TABLE 1 Inclusion and Exclusion criteria**

INCLUSION CRITERIA

All participants will be aged 16 years or over

Participants must be resident in the United Kingdom

EXCLUSION CRITERIA

The inability to provide informed consent

Under the age of 16 years

A non-UK resident

Kidney disease

High levels of calcium in the blood

Atherosclerosis

Sarcoidosis

Histoplasmosis

Over-active parathyroid gland (hyperparathyroidism)

Lymphoma

Currently taking thiazide diuretics, digoxin or other cardiac glycosides

Known allergy to nuts (peanut oil contained within vitamin D preparations)

Female subjects of child bearing age

Established on vitamin D or multivitamin supplementation

**SUPPLEMENTARY FIGURE 1 Study flow diagram**

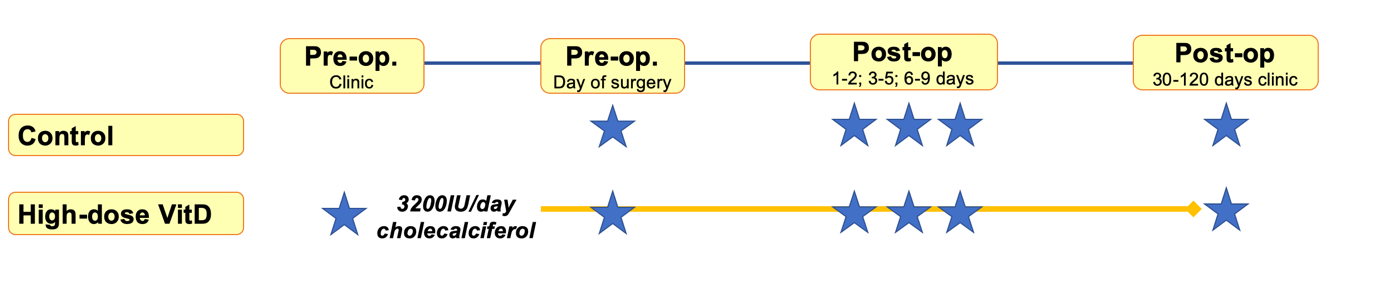

** indicates sample point*

**SUPPLEMENTARY TABLE 2 Clinicopathological demographics of included patients stratified by control group sampling years**

|  | **Control 2016** | **Control 2020** | **P** |
| --- | --- | --- | --- |
| N | 62 | 19 | - |
| Sampling Years | 2013 | 2020 | - |
| Age (mean) | 67.5 | 60.0 | 0.14 |
| Gender (F) | 29 (47%) | 8 (42%) | 0.80 |
| BMI (kg/m^2^) | 27.24 | 27.20 | 0.59 |
| Baseline 25OHD (mean, 95%CI)) | 46.3  (39.8-52.9) | 47.6  (34.9-60.3) | 0.86* |
| Cancer site |  |  | - |
| Colon | 39 (63%) | 10 (53%) | 0.43 |
| Rectum | 23 (37%) | 9 (47%) | 0.43 |
| Cancer stage |  |  | - |
| AJCC 1 | 19 (31%) | 7 (37%) | - |
| AJCC 2 | 25 (40%) | 2 (11%) | - |
| AJCC 3 | 12 (19%) | 7 (37%) | - |
| AJCC 4 | 6 (10%) | 3 (16%) | *Ptrend=0.44* |

**P value for 25OHD comparison between control groups using fixed effects model adjusting for gender, AGE, BMI and AJCC P=0.72*

**SUPPLEMENTARY TABLE 3 Full multivariate mixed-effects model for factors associated with peri-operative 25OHD level.**

| Factor | Estimate | P |
| --- | --- | --- |
| Treatment group |  |  |
| Control | ref | ref |
| High-dose VitD | 62.2 | 1.22E-15 |
| Sample point |  |  |
| Pre-op | ref | ref |
| 1-2 days | -18.1 | 0.04 |
| 3-5 days | -12.4 | 0.28 |
| 6-9 days | -23.4 | 0.01 |
| Clinicodemographic factors |  |  |
| Age | 0.1 | 0.67 |
| Gender (M) | -2.1 | 0.44 |
| BMI | -1.0 | 0.02 |
| AJCC | -1.3 | 0.48 |

*P value given adjusted in fixed effects model for gender, AGE, BMI and AJCC. Estimate derived from model run before log transformation to provide raw estimate of effect of each factor on 25OHD level. When control groups were considered individually, significant impact of supplementation remained (2016 control group estimate ~63nmol/l, P=1.9E-15; 2020 control group estimate ~60nmol/l, P=3.51E-08).*

**SUPPLEMENTARY TABLE 4 Post-operative 25OHD fold-change compared to day of surgery pre-operative level**

|  | Control | High-dose VitD | |
| --- | --- | --- | --- |
| Timepoint | **FC (iqr)** | **FC (iqr)** | **P** |
| Day 1-2 | 0.54 (0.38) | 0.76 (0.27) | 0.0003 |
| Day 3-5 | 0.56 (0.44) | 0.9 (0.29) | 0.0001 |
| Day 6-9 | 0.63 (0.42) | 0.78 (0.59) | 0.17 |

*Median fold-change in unadjusted-adjusted 25OHD given (iqr)*

**SUPPLEMENTARY FIGURE 2 Peri-operative CRP levels**

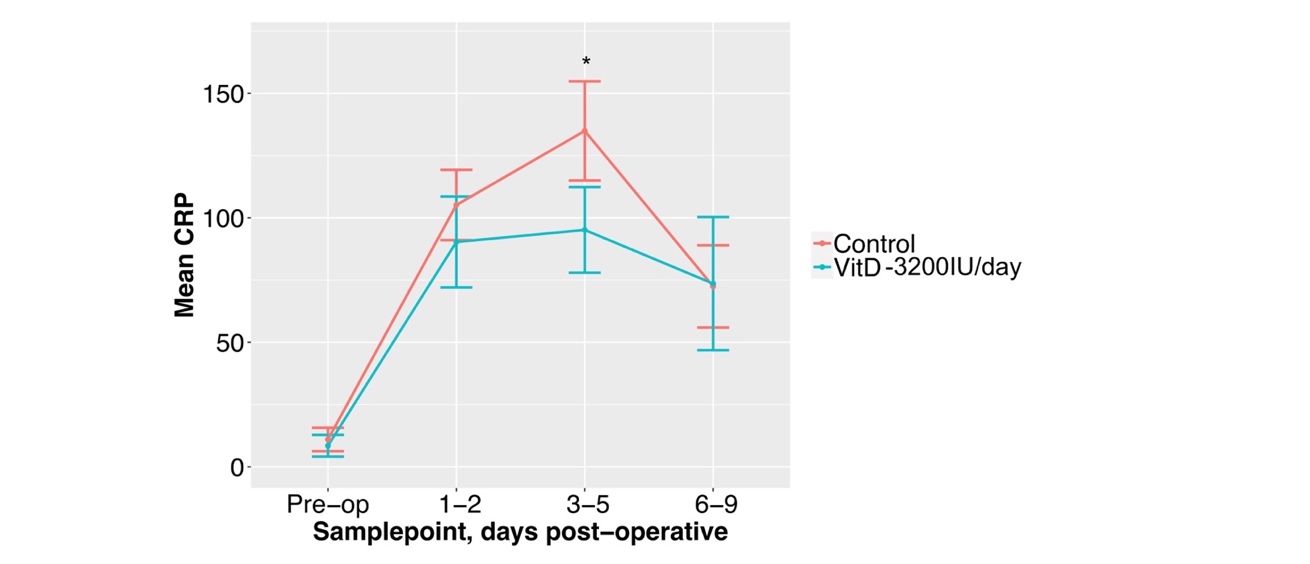

**SUPPLEMENTARY TABLE 5 Full multivariate mixed-effects model for factors associated with early post-operative CRP level**

| Factor | Estimate | P |
| --- | --- | --- |
| Treatment group |  |  |
| Control | ref | ref |
| High-dose VitD | -2.65 | 0.035 |
| Sample point |  |  |
| Pre-op | Ref | ref |
| 1-2 days | 81 | <2.00E-16 |
| 3-5 days | 87 | <2.00E-16 |
| 6-9 days | 63 | <2.00E-16 |
| Clinicodemographic factors |  |  |
| Age | -0.16 | 0.047 |
| Gender (M) | 28.22 | 0.009 |
| BMI | -0.50 | 0.03 |
| AJCC | 9.51 | 0.14 |
